# Allostatic load as a potential pathway linking exposomic risk to early-onset cancer: a matched case-control study in the All of Us Research Program

**DOI:** 10.64898/2026.10.31.26366980

**Authors:** Ravi B. Parikh, Anthony Girard, Tyler M. Moore, Elina Visoki, Timothy Bigdeli, Sinan Guloksuz, Lauren E. McCullough, Katherine L. Nathanson, Ran Barzilay

**Affiliations:** Emory University, Atlanta, GA, USA; University of Pennsylvania, Philadelphia, PA, USA; Children’s Hospital of Philadelphia, Philadelphia, PA, USA; SUNY Downstate Health Sciences University, Brooklyn, NY, USA; Maastricht University, Maastricht, the Netherlands; Rollins School of Public Health, Emory University, Atlanta, GA, USA; Division of Translational Medicine and Human Genetics, Department of Medicine, Perelman School of Medicine, University of Pennsylvania, Philadelphia, PA, USA

**Keywords:** early-onset cancer, allostatic load, exposome, mediation analysis, social determinants of health, All of Us

## Abstract

**Background:** Early-onset cancer (EoC) incidence is rising worldwide, yet risk models built on isolated exposures or rare pathogenic variants capture little of the cumulative environmental burden experienced by younger adults. Allostatic load (AL), the cumulative physiological burden of chronic stress, can be measured from routine laboratory values and vital signs and is a candidate biological pathway linking environmental exposures to carcinogenesis.

**Methods:** We conducted a 1:4 matched case-control study nested within the NIH All of Us Research Program, including 7,550 participants and 1,510 cancers diagnosed before age 50, of which 298 were exposome-related (the primary outcome). We derived an exposomic risk score (ERS) from 12 survey-based measures and an eight-component AL score from electronic health record data, and used mediation analysis to estimate the proportion of the ERS-EoC association operating through AL.

**Results:** A higher ERS was associated with higher AL (β=0.18 standard deviations [SD] per SD; 95% CI 0.16-0.20), and each SD increase in AL was associated with 21% higher odds of EoC (odds ratio 1.21; 95% CI 1.15-1.28). For exposome-related EoC, AL mediated 17.4% (95% CI 8.2-44.7%) of the association between the ERS and EoC, with significant total, direct, and indirect effects that were robust to sensitivity analyses of diagnosis timing, AL scoring, and age adjustment. Across all EoC, the proportion mediated was 48.8% (95% CI 21.2-186.5%).

**Conclusions:** AL is a candidate biomarker and potential pathway through which cumulative environmental burden relates to EoC, warranting prospective validation in independent cohorts.

## Introduction

Early-onset cancer (EoC), defined as cancer diagnosed before age 50, is an escalating public health concern that challenges conventional paradigms of cancer risk assessment. Globally, the number of incident EoC cases increased by 79.1% between 1990 and 2019,^1^ and in the United States gastrointestinal cancers have shown the fastest-growing incidence among adults younger than 50.^2,3^ Increased imaging and screening may explain part of this trend,^4^ but increases have also been observed for malignancies without routine screening in younger adults, such as pancreatic cancer, suggesting factors beyond enhanced detection.^2,5^ The economic burden is substantial, with an estimated $150 billion in annual US productivity losses attributed to premature mortality, fertility preservation, and caregiver burden.^6,7^ Current prevention strategies, including the lowering of the recommended starting age for colorectal cancer screening, may inadequately address the heterogeneous risk factors underlying EoC while placing growing demands on screening capacity such as colonoscopy.^8,9^

Traditional cancer risk assessment paradigms, largely developed in older populations, rely on identifying individuals with isolated environmental exposures (for example, smoking, alcohol, or a low-fiber diet) or high-penetrance pathogenic germline variants (for example, Lynch syndrome).^10–12^ Single-exposure models do not capture the co-occurring, cumulative nature of EoC risk factors, and hereditary cancer syndromes account for a minority of EoC cases.^13–15^ The exposome framework, first articulated as a complement to the genome,^16^ considers the totality of exposures across the life course, including environmental pollutants, lifestyle and psychosocial factors, and the physiological responses they elicit.^17,18^ By accounting for co-occurring exposures, this approach may improve cancer risk stratification relative to single-exposure studies, particularly for EoC, where individual exposures have shown limited explanatory power.^19–21^

Allostatic load (AL), the cumulative physiological burden resulting from chronic adaptation to stress, offers a framework for understanding how environmental exposures may translate into disease risk.^22,23^ AL represents “wear and tear” across neuroendocrine, immune, metabolic, and cardiovascular systems and is associated with adverse mental and physical health outcomes, all-cause mortality, and accelerated biological aging.^24–26^ AL partially mediates associations between environmental or genetic factors and other conditions that arise in younger people, including youth mental health conditions, preterm birth, and early-onset cardiovascular disease.^27–30^ In cancer, AL has been associated with all-cause and cancer-specific mortality and, in older adults, with colorectal cancer incidence (Supplementary Table 2).^31–34^ Whether AL lies on the pathway between cumulative environmental burden and EoC has not been examined, despite growing exposure among young adults to chronic stressors such as economic uncertainty, job insecurity, and housing unaffordability.^35–37^

We hypothesized that AL is a quantifiable, potentially modifiable pathway through which exposomic burden relates to EoC. The physiological systems that compose AL (metabolic, cardiovascular, immune, and neuroendocrine) also influence cancer biology through chronic inflammation, metabolic dysfunction, impaired immune surveillance, and genomic instability (Supplementary Table 1). To test this hypothesis, we conducted a matched case-control study nested within the NIH All of Us Research Program (AoU), comprising 1,510 cancers diagnosed before age 50. We derived an exposomic risk score capturing multi-domain environmental burden, quantified AL from routinely collected clinical measures, and used mediation analysis to estimate the extent to which AL accounts for the association between exposomic burden and EoC. The primary outcome was exposome-related EoC, comprising cancers with established environmental and lifestyle risk factors.

## Results

### Study design and population

The analysis proceeded in three phases (Fig. 1): quantification of AL from clinical measures, derivation of an exposomic risk score (ERS) from survey data, and mediation analysis testing AL as a pathway linking the ERS to EoC. We identified 1,510 AoU participants with a first cancer diagnosed before age 50 after enrollment, excluding breast cancer (Methods), and matched each 1:4 to participants with no cancer diagnosis at any time, exactly on age and sex and by propensity score on sociodemographic characteristics. The analytic sample comprised 7,550 participants (1,510 cases and 6,040 controls). Cases and controls were balanced on age (mean 39.3 years, SD 7.53), sex (73.4% female), race and ethnicity, education, and income (Table 1). Of the 1,510 cases, 298 had exposome-related cancers (head and neck, digestive, respiratory, soft tissue, and urinary tract; Supplementary Table 6); these formed the primary outcome and were compared with all 6,040 controls. Exposome measures were obtained from surveys completed at enrollment. AL components were obtained from electronic health record (EHR) data recorded 1-3 years before diagnosis for cases and within the 2 years before enrollment for controls (Supplementary Fig. 1). Cases were diagnosed a median of 1.5 years (IQR 0.7-2.7) after enrollment.

**Figure 1.**
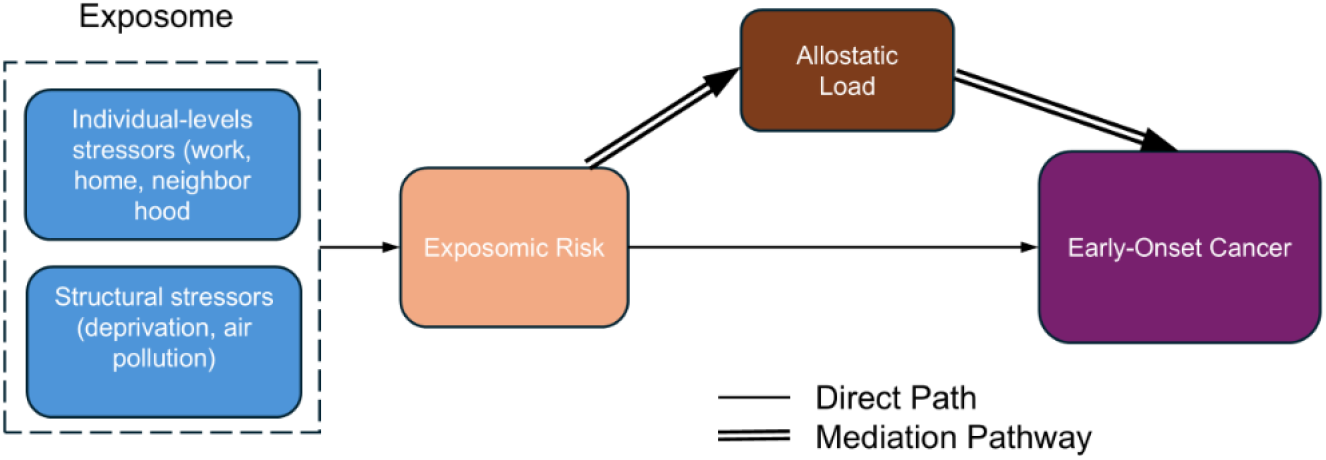
Conceptual framework. Hypothesized pathway from exposomic burden through allostatic load to early-onset cancer. Double lines indicate the mediated (indirect) pathway; the single line indicates the direct pathway. The exposome is depicted conceptually; the domains measured in this study are listed in Supplementary Table 3 and did not include ambient environmental exposures such as air pollution.

**Table 1.** Baseline characteristics of matched cases and controls.

| **Characteristic** | **Cases (n=1,510)** | **Controls (n=6,040)** |
|---|---|---|
| Female, n (%) | 1,108 (73.4) | 4,432 (73.4) |
| Age at enrollment, mean (SD), years | 39.3 (7.53) | 39.3 (7.53) |
| **Race and ethnicity, n (%)** |  |  |
| Asian | 40 (2.6) | 134 (2.2) |
| Black | 265 (17.5) | 1,090 (18.0) |
| Hispanic | 317 (21.0) | 1,274 (21.1) |
| White | 833 (55.2) | 3,337 (55.2) |
| Other | 55 (3.6) | 205 (3.4) |
| **Education, n (%)** |  |  |
| High school | 281 (18.6) | 1,140 (18.9) |
| Some college | 432 (28.6) | 1,748 (28.9) |
| College degree | 651 (43.1) | 2,609 (43.2) |
| Unspecified | 146 (9.7) | 543 (9.0) |
| **Annual household income, n (%)** |  |  |
| <\$50,000 | 659 (43.6) | 2,575 (42.6) |
| \$50,000-100,000 | 273 (18.1) | 1,117 (18.5) |
| >\$100,000 | 349 (23.1) | 1,454 (24.1) |
| Unspecified | 229 (15.2) | 894 (14.8) |

Cases and controls were matched 1:4, exactly on age at enrollment and sex, and by propensity score on race, insurance status, employment status, education, and income. Cases were participants diagnosed with a first cancer (excluding breast cancer) before age 50 after enrollment.

### Allostatic load

AL was defined as the number of eight components outside clinical reference ranges, using the most recent value in each participant’s measurement window, and was standardized for analysis. The components were blood pressure, body mass index (BMI), glucose, hemoglobin, white blood cell count (WBC), albumin, creatinine, and calcium (Supplementary Table 4). The mean AL count was 0.78 (SD 1.05), and 19.6% of participants had two or more abnormal components. AL increased modestly with age (r=0.07; p<0.001) and was higher in female than in male participants (mean 0.80 vs 0.73; p=0.020). Overall, 30.1% of participants had all eight components measured; albumin had the highest missingness (56.9%) (Supplementary Table 5). Missing components were imputed using random forests (Methods).

### Allostatic load and early-onset cancer

Each SD increase in AL was associated with higher odds of EoC (odds ratio [OR] 1.21, 95% CI 1.15-1.28; p<0.001; Fig. 2) and exposome-related EoC (OR 1.32, 95% CI 1.20-1.45; p<0.001) after adjustment for age, sex, race and ethnicity, income, and education. Associations were also observed using a binary indicator of two or more abnormal components (OR 1.54, 95% CI 1.35-1.76) and a factor-analytic AL score (OR 1.26, 95% CI 1.18-1.35) (Table 2). In a complete-case analysis restricted to participants with all eight components measured (n=2,272), the association was directionally consistent with the primary analysis but attenuated and not statistically significant (OR 1.06, 95% CI 0.98-1.14; p=0.125). The proportion of participants who were cases increased from 17.1% among those with no abnormal components to 27.3% among those with three (Fig. 3).

**Figure 2.**
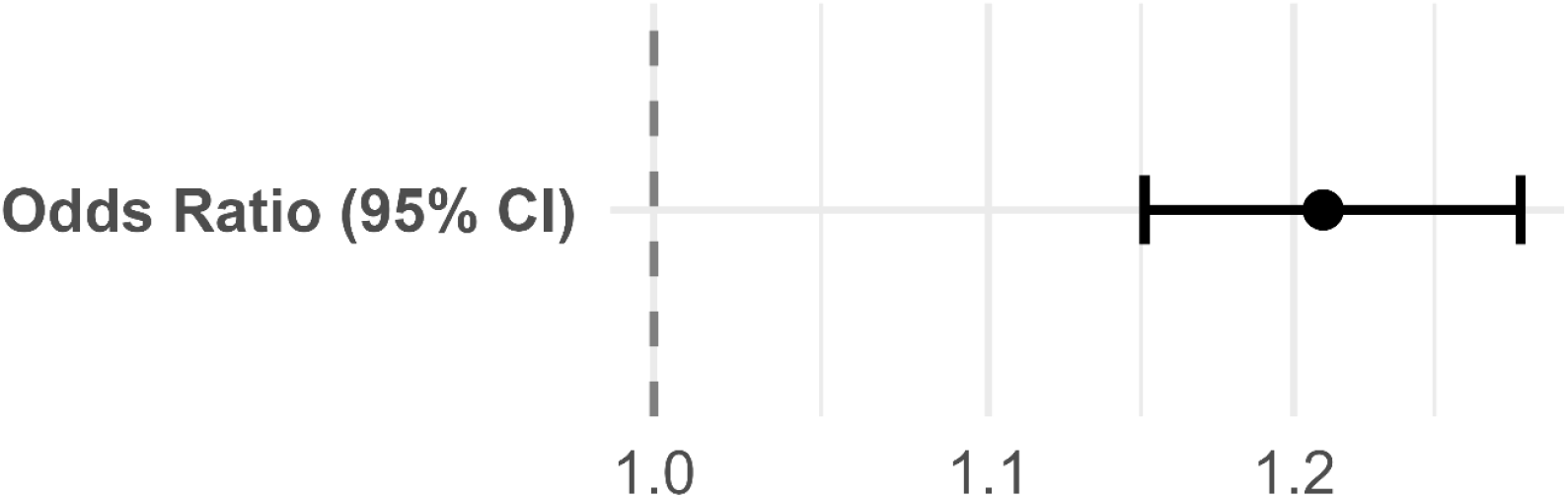
Association between allostatic load and early-onset cancer. Odds ratio and 95% confidence interval per 1-SD increase in the primary AL score, from logistic regression of all early-onset cancer adjusted for age, sex, race and ethnicity, income, and education.

**Figure 3.**
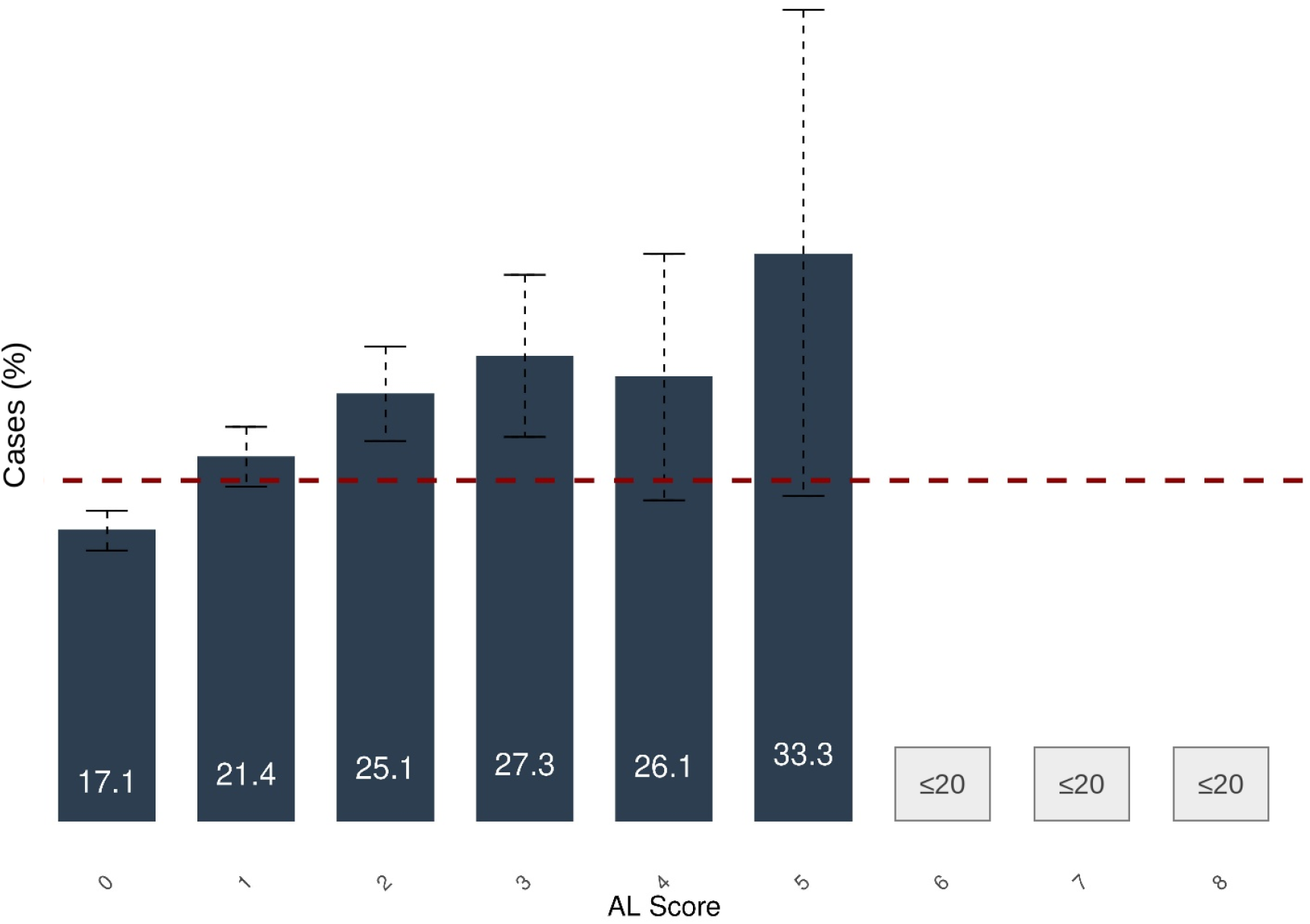
Proportion of participants who were early-onset cancer cases, by allostatic load score. Bars show the percentage of participants at each AL score (count of abnormal components, 0-8) who were cases; error bars show 95% confidence intervals. The dashed line indicates the overall case proportion of 20% fixed by 1:4 matching. Boxes marked ≤20 indicate AL scores with 20 or fewer participants or cases, which are suppressed in accordance with the AoU Data and Statistics Dissemination Policy.

**Table 2.** Association between allostatic load and early-onset cancer by AL scoring method.

| AL score | Data | Odds ratio (95% CI) | p |
|---|---|---|---|
| Primary (standardized count of abnormal components) | Imputed | 1.21 (1.15-1.28) | <0.001 |
| Binary (≥2 vs <2 abnormal components) | Imputed | 1.54 (1.35-1.76) | <0.001 |
| Factor-analytic score | Imputed | 1.26 (1.18-1.35) | <0.001 |
| Primary (standardized count of abnormal components) | Complete case (n=2,272) | 1.06 (0.98-1.14) | 0.125 |

Odds ratios for all early-onset cancer from logistic regression adjusted for age, sex, race and ethnicity, income, and education; per 1-SD increase for continuous scores and high versus low for the binary score. Imputed analyses used random-forest imputation (missForest) of missing AL components; the complete-case analysis was restricted to participants with all eight components measured. AL, allostatic load; CI, confidence interval.

### Exposomic risk score

The ERS was derived from 12 survey-based measures, eight of which were validated multi-item scales, spanning neighborhood environment (walkability, crime and safety, social cohesion, and social and physical disorder), discrimination (everyday and in health care settings), social support, household context (income, household size, and housing instability), and substance use (Supplementary Table 3). Parallel analysis supported a three-component solution with bifactor rotation, which explained 52.6% of the total variance. Loadings on the general factor ranged from −0.79 to 0.70; the strongest were for social support (−0.79), discrimination in health care settings (0.70), and neighborhood crime and safety (0.70), such that higher scores reflect greater adverse exposure. Two group factors captured neighborhood environment and household context and discrimination; these were not used in primary analyses. The standardized general factor served as the ERS.

### Exposomic risk and allostatic load

In a univariate analysis, higher ERS was associated with higher AL (β=0.18 SD per SD of ERS, 95% CI 0.16-0.20; p<0.001)

### Allostatic load as a potential pathway between exposomic risk and early-onset cancer

For the primary outcome, exposome-related EoC (298 cases and 6,040 controls), the ERS was associated with case status (total effect 0.018, 95% CI 0.008-0.028; p<0.001), with significant direct effects (0.014, 95% CI 0.004-0.024; p=0.004) and indirect effects through AL (0.003, 95% CI 0.002-0.004; p<0.001). AL mediated 17.4% (95% CI 8.2-44.7%) of the total effect, consistent with partial mediation (Table 3). For all EoC, the total effect was 0.022 (95% CI 0.005-0.040; p=0.016) and the indirect effect 0.011 (95% CI 0.008-0.013; p<0.001); the direct effect was not significant (0.011, 95% CI −0.006 to 0.030; p=0.210), and the proportion mediated was 48.8% (95% CI 21.2-186.5%). Effects are differences in the predicted probability of case status comparing an ERS 1 SD above versus 1 SD below the mean. Because cases made up about 5% of the exposome-related analysis and 20% of the all-EoC analysis, effects on this scale are not directly comparable between the two outcomes (Methods).

**Table 3.**
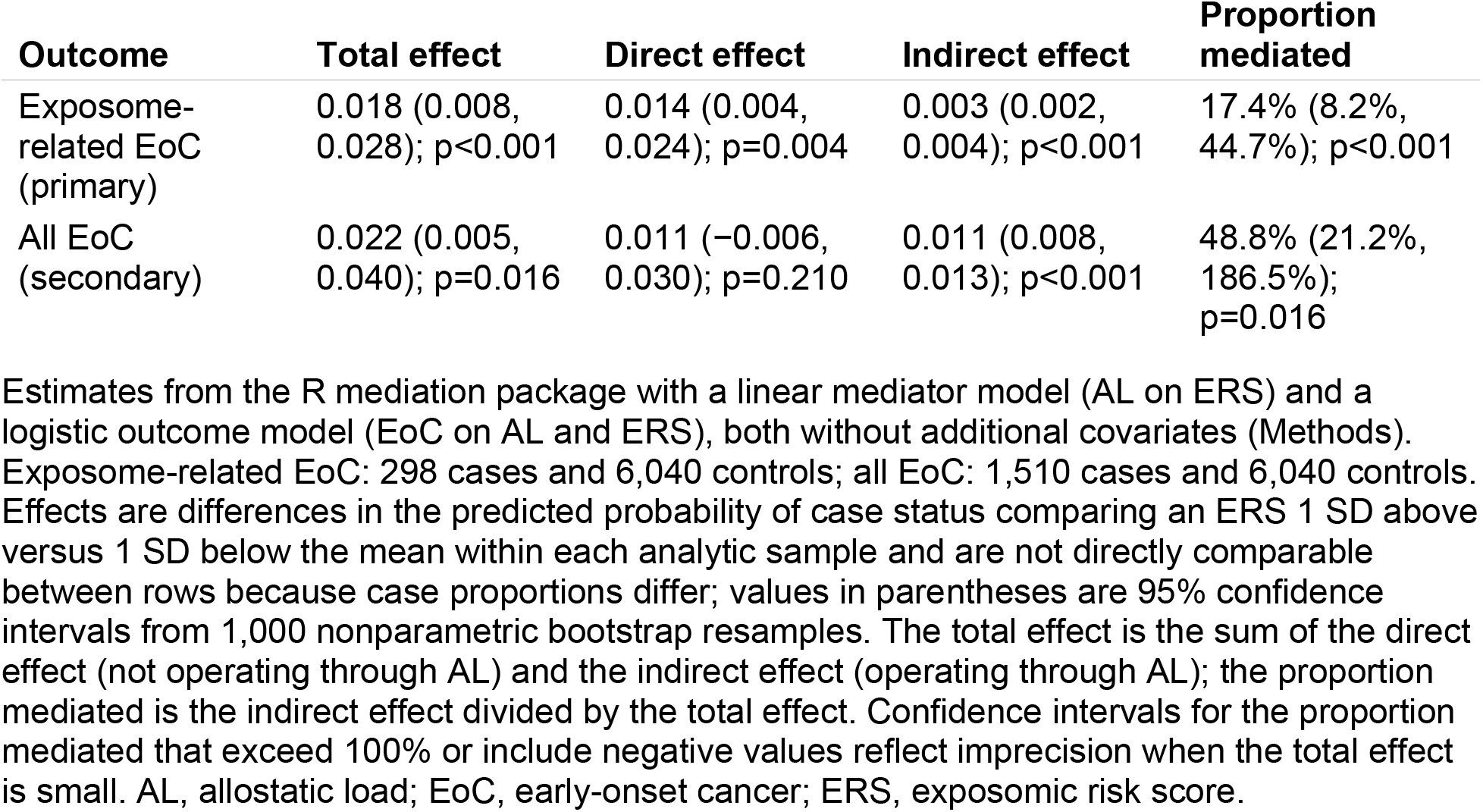
Mediation of the association between exposomic risk and early-onset cancer by allostatic load.

### Sensitivity analyses

For exposome-related EoC, the proportion mediated was stable across sensitivity analyses (Supplementary Table 8): 16.4% when restricted to cases diagnosed at least 2 years after enrollment, 16.0% after adding age to both models, 12.0% using the factor-analytic AL score, and 14.8% using the binary AL score. In the restricted analysis, effects for exposome-related EoC were attenuated but remained significant (total effect 0.008, p=0.010; indirect effect 0.001, p<0.001). For all EoC, this restriction reduced the indirect effect from 0.011 to 0.004 and the proportion mediated from 48.8% to 21.8%, although both total and indirect effects remained significant. In sex-stratified analyses, AL mediated 11.5% (95% CI 4.2-42.2%) of the association with exposome-related EoC among male participants; among female participants, the indirect effect was significant (p<0.001) but the total effect was not (p=0.114). In the complete-case analysis, indirect effects were directionally consistent with the primary analysis but not statistically significant (exposome-related EoC: 0.002, 95% CI −0.002 to 0.006).

Generalized additive models showed modest nonlinearity in the ERS-AL association (effective degrees of freedom [edf] 5.31; p<0.001 versus linear), although the smooth term reduced deviance by less than 1%; AL increased with ERS up to approximately 2 SD above the mean and attenuated in the upper tail (Supplementary Fig. 2a). The AL-EoC association showed modest departure from linearity on the log-odds scale (edf 2.04; p=0.028), with a monotonic increase and slight attenuation at higher AL values (Supplementary Fig. 2b). Linear specifications were therefore retained as parsimonious summaries. Replacing the linear mediator model with a generalized additive model yielded the same pattern of results, with somewhat larger indirect effects: AL mediated 24.1% (95% CI 13.3-54.0%) of the association with exposome-related EoC and 55.7% (95% CI 28.7-209.4%) of the association with all EoC (Supplementary Table 8).

### Exploratory analyses by cancer type

Associations between AL and EoC varied by cancer type (Supplementary Table 7). Odds ratios exceeded 1 for all five exposome-related cancer groups, ranging from 1.18 (respiratory) to 1.50 (urinary tract), and the confidence interval excluded 1 for urinary tract (OR 1.50, 95% CI 1.26-1.77), head and neck (OR 1.45, 95% CI 1.10-1.91), and digestive cancers (OR 1.24, 95% CI 1.07-1.44), the largest exposome-related group. Outside this group, the strongest association was for hematologic and lymphoid malignancies (OR 1.46, 95% CI 1.34-1.58), which may partly reflect pre-diagnostic changes in WBC and hemoglobin (Discussion); an association was also observed for female genital cancers (OR 1.16, 95% CI 1.04-1.30). Skin cancers, which included keratinocyte carcinomas (C44), showed an inverse association (OR 0.83, 95% CI 0.70-0.99), which may reflect health care utilization patterns, such as dermatologic surveillance, rather than biology. These analyses were not powered to detect associations of the magnitude observed overall and should be considered exploratory.

## Discussion

In this matched case-control study within the All of Us Research Program, a cumulative exposomic risk score was associated with higher allostatic load, AL was associated with higher odds of early-onset cancer, and AL accounted for part of the association between exposomic burden and EoC. For the primary outcome of exposome-related EoC, total, direct, and indirect effects were all significant, indicating partial mediation, with approximately one-sixth of the association operating through AL. These findings are consistent with the hypothesis that AL is a quantifiable physiological pathway through which cumulative environmental exposures relate to cancer in younger adults, and they suggest potential roles for AL in risk stratification and, if the pathway is causal, in prevention.

Environmental exposures contribute substantially to cancer burden, yet risk assessment paradigms that examine single exposures do not capture the cumulative, co-occurring nature of environmental risk factors underlying EoC. Our approach addresses this limitation by deriving an integrated score from 12 survey measures using principal components analysis with bifactor rotation, eight of them validated multi-item instruments, spanning neighborhood environment, discrimination, social support, household context, and substance use. The association between the ERS and AL (β=0.18) aligns with literature linking adverse social and environmental conditions to cardiometabolic and immune dysregulation.^38,39^ Prior work has linked isolated exposures, such as smoking, alcohol consumption, and neighborhood deprivation, to EoC^40–42^; our findings extend this work by showing that cumulative psychosocial and neighborhood burden is associated with measurable physiological dysregulation that is, in turn, associated with EoC.

A key finding is that the AL pathway was most consistent for exposome-related cancers, including digestive, respiratory, urinary tract, and head and neck malignancies. For these cancers, AL mediated 17.4% of the association between exposomic burden and EoC; this proportion was stable across diagnosis-timing, AL scoring, and age-adjusted analyses (12.0-17.4%) and somewhat higher with a nonlinear mediator model (24.1%). By contrast, the larger proportion mediated for all EoC (48.8%) was imprecise and fell to 21.8% when the analysis was restricted to cases diagnosed at least 2 years after enrollment. This pattern aligns with mechanistic evidence linking AL to carcinogenesis through chronic inflammation, metabolic dysfunction, immune suppression, and genomic instability.^43–45^ Sustained activation of stress pathways may elevate pro-inflammatory cytokines that create tumor-promoting microenvironments, while metabolic dysregulation may alter cellular energetics in ways that favor malignant transformation.^46,47^ Our findings extend prior evidence of AL’s mediating role in other conditions of younger adults, including youth mental health conditions, preterm birth, and early-onset cardiovascular disease,^27–30^ to EoC, suggesting that the physiological embedding of chronic environmental stress may be one mechanism underlying EoC and a potentially modifiable target.

Although we did not incorporate inherited genetic factors, germline risk does not account for the majority of EoC. High-penetrance syndromes such as Lynch syndrome and pathogenic variants in *BRCA1* and *BRCA2* explain a minority of cases,^13,14^ established risk prediction models perform less well among individuals younger than 50,^11,48^ and many patients with EoC present without traditional risk factors.^40,49^ The extent to which aggregate polygenic risk, which indexes both direct cancer susceptibility and indirect risk through liability to predisposing traits, contributes to EoC relative to environmental factors remains unclear. In prior work in a healthy adolescent sample, we observed gene-environment interactions in association with AL, in which polygenic risk for metabolic and stress-related conditions amplified the association between exposomic burden and AL.^27^ Future EoC studies that integrate polygenic risk scores and pathogenic germline variants with exposomic and AL measures could clarify whether genetic predisposition modifies EoC risk through AL-related mechanisms.

Exploratory cancer-type analyses suggested heterogeneity in the AL-EoC association, with the strongest association for hematologic malignancies. These analyses were limited by small numbers of cases for individual cancer types, and associations for cancers that directly perturb AL components should be interpreted cautiously. Larger cohorts will be needed to determine whether AL-related pathways differ by tumor site and molecular subtype.

These findings should be interpreted in the context of several limitations. First, AL comprised eight clinical measures available in EHR data; HbA1c and lipids were excluded because of high missingness, and key stress mediators, including cortisol, dehydroepiandrosterone sulfate, and inflammatory markers such as C-reactive protein, were unavailable, which may have attenuated associations. Second, missingness in AL components was substantial (69.9% of participants had at least one missing component). Complete-case estimates were directionally consistent with the primary analysis but attenuated and not statistically significant, so our findings depend in part on imputation. Because random-forest imputation tends to predict values within reference ranges, imputed components were rarely classified as abnormal; if the number of measured components differed between cases and controls, this could bias associations. Third, AL measurement windows were anchored to diagnosis for cases and to enrollment for controls. Because cases were diagnosed a median of 1.5 years after enrollment and the most recent value in each window was used, case AL values were often recorded after the exposome survey and closer to the outcome, whereas control values always preceded enrollment. This asymmetry could bias the AL-EoC association, and it means the order in which exposure and mediator were measured differed between groups. Fourth, although AL was measured at least 1 year before diagnosis, we cannot exclude reverse causation, and we did not test longer intervals between AL measurement and diagnosis. Restricting to cases diagnosed at least 2 years after enrollment, which lengthened the interval between exposome assessment and diagnosis, left the proportion mediated for exposome-related EoC similar but more than halved it for all EoC. Relatedly, several AL components (hemoglobin, WBC, creatinine, and calcium) can be altered by undiagnosed malignancy (for example, anemia preceding gastrointestinal or gynecologic cancers, or cytopenias preceding hematologic malignancies), which may inflate associations, particularly for all EoC. Fifth, mediation models were not adjusted for covariates. Matching balanced cases and controls on sociodemographic factors, but it does not address confounding of the exposome-AL association, and for the primary outcome, exposome-related cases were compared with controls matched to the full case series. Adding age to both models produced similar estimates, but residual confounding by sex, race and ethnicity, or education cannot be excluded. Sixth, because cases and controls were sampled on outcome status, mediation effects on the probability scale reflect the case-control sample rather than population risks, the mediator model was estimated in a case-enriched sample, and effects are not directly comparable between outcomes with different case proportions; these estimates are best interpreted as measures of the relative contribution of the AL pathway rather than as absolute risk differences. Seventh, household income contributed to the ERS and to the propensity score used for matching, which may have attenuated the total effect of the ERS. Eighth, the ERS was limited to continuous survey measures; binary exposures and several domains, including diet, physical activity, early-life adversity, and ambient environmental exposures such as air pollution, were not represented. Finally, cancer outcomes were ascertained from EHR data rather than registry linkage; the secondary all-EoC outcome included keratinocyte carcinomas, diagnoses without an assignable site, and a small number of in situ diagnoses; breast cancer was excluded; and AoU participants with EHR and laboratory data may differ from the general population of younger adults.

In conclusion, in this matched case-control study within a large, diverse US cohort, allostatic load was associated with early-onset cancer and partially accounted for the association between cumulative exposomic burden and EoC. These findings support further study of AL as a candidate biomarker for identifying younger adults whose environmental burden has translated into measurable physiological dysregulation. If confirmed prospectively and in independent cohorts, behavioral, pharmacological, or policy interventions targeting AL components may represent strategies for reducing EoC risk, particularly for exposome-related cancers, for which the AL pathway was most consistent across analyses. Future research should integrate polygenic risk, pathogenic germline variants, and longitudinal exposome assessments to refine mechanistic understanding and enable precision prevention.

## Methods

### Study design

We conducted a 1:4 matched case-control study nested within the NIH All of Us Research Program, with exposomic measures and AL components ascertained before cancer diagnosis. The analysis followed three phases (Fig. 1). Phase 1 quantified AL from a predefined set of clinical measures capturing cumulative physiological stress.^31^ Phase 2 quantified exposomic burden from survey measures, reduced through bifactor analysis to a single exposomic risk score (ERS). Phase 3 applied mediation analysis to estimate the extent to which AL accounts for the association between the ERS and EoC. Controls were selected from participants who remained cancer-free through the end of available follow-up (cumulative rather than risk-set sampling); odds ratios therefore approximate relative risks only under the rare-disease assumption. This study is reported in accordance with the STROBE guideline for case-control studies (Supplementary Information).

### Data source

The All of Us Research Program is a national, population-based cohort led by the US National Institutes of Health that aims to enroll at least one million adults aged 18 years and older, with intentional inclusion of populations historically underrepresented in biomedical research.^50^ Enrollment began in 2018 and includes baseline surveys, physical measurements, and biospecimens, complemented by retrospective and prospective EHR linkage, follow-up surveys, and optional digital health data. As of late 2025, more than 870,000 individuals had enrolled, with almost 630,000 contributing baseline data.^51^ This study used the All of Us Controlled Tier Dataset v9.

### Participants

Among more than 393,000 AoU participants with available EHR data, 68,833 had a cancer diagnosis recorded in the EHR. We excluded participants whose first cancer diagnosis was breast cancer (International Classification of Diseases, Tenth Revision [ICD-10] C50; n=11,088), leaving 57,745. Breast cancer was excluded because its etiology is predominantly hormone-mediated (driven by estrogen and progesterone receptor signaling and by reproductive factors) rather than driven by the cardiometabolic, inflammatory, and stress-response pathways that AL captures. Its inclusion would be expected to obscure AL-related associations with other cancers, and because breast cancer is the most common cancer among women younger than 50, it would otherwise have accounted for a large share of cases. Restricting to participants diagnosed before age 50 yielded 13,362 (n=44,383 excluded), of whom 1,906 were diagnosed after AoU enrollment (n=11,456 excluded); participants diagnosed before enrollment were not eligible. Requiring at least one measured AL component yielded 1,510 cases (n=396 excluded). No exclusion based on time from enrollment to diagnosis was applied; instead, the AL window for cases ended 1 year before diagnosis (see Temporal framework).

Controls were AoU participants younger than 50 at enrollment with no cancer diagnosis recorded at any time and at least one measured AL component. Cases were matched to controls 1:4 without replacement. Age at enrollment and sex were matched exactly, and within these strata, nearest-neighbor matching was performed on a propensity score estimated by logistic regression including age, sex, race, insurance status, employment status, education, and income. The final analytic sample comprised 7,550 participants (1,510 cases and 6,040 controls).

### Temporal framework

Exposomic measures were derived from surveys completed at enrollment. For controls, AL components were obtained from EHR data recorded within the 2 years before consent to EHR sharing at enrollment. For cases, AL components were obtained from EHR data recorded 1-3 years before cancer diagnosis, so that at least 1 year separated AL measurement from diagnosis; biomarker values recorded shortly before diagnosis may reflect undiagnosed disease or diagnostic workup.^52^ Within each window, the most recent value of each component was used, and multiple values recorded on the same day were averaged. Supplementary Fig. 1 illustrates the temporal framework. Because each participant was anchored to a fixed index date rather than followed for time to event, time from enrollment to diagnosis among cases is reported descriptively.

### Exposomic risk score

Exposome variables were derived from AoU surveys in the following domains: The Basics, Overall Health, Lifestyle, Personal and Family Health History, Health Care Access and Utilization, and Social Determinants of Health (SDoH). Although SDoH items are stored as individual questions, they derive from validated instruments designed to be analyzed as multi-item scales; we therefore reconstructed each scale and computed summary scores using the published scoring methods. Summary scores were calculated as the mean of answered items, allowing zero, one, or two missing items depending on the instrument; scores were set to missing when this threshold was exceeded. Of 21 exposome-related variables identified, nine binary items were excluded from the factor analysis because their inclusion produced an unstable factor solution; they were retained as predictors in the imputation model. The final set comprised 12 variables (Supplementary Table 3).

Missing exposome values were imputed with random forests implemented in the h2o package.^53,54^ For each variable with missing data, a random forest was trained using all other variables as predictors and used to predict the missing values; variables were imputed sequentially in randomized order, with previously imputed values used as predictors for subsequent variables. Continuous and categorical variables were imputed using regression and classification forests, respectively.

We applied principal components analysis with bifactor rotation,^55^ in which all variables load on a general component and on orthogonal domain-specific group components; consistent with prior work, we refer to the general component as the general exposome factor.^56^ The number of components was selected by parallel analysis, which supported three (one general and two group components) because the first three observed eigenvalues exceeded those from simulated data. Scores on the general component, interpretable as overall exposomic burden, were standardized to mean 0 and SD 1 to form the ERS. Analyses used the psych package in R.^57^

### Allostatic load

AL was originally quantified using ten biomarkers summarizing cardiovascular, metabolic, hypothalamic-pituitary-adrenal axis, and sympathetic nervous system activity: systolic and diastolic blood pressure, waist-to-hip ratio, high-density lipoprotein and total cholesterol, glycated hemoglobin (HbA1c), serum dehydroepiandrosterone sulfate, and 12-hour urinary cortisol, norepinephrine, and epinephrine.^58,59^ Subsequent operationalizations frequently incorporate additional markers such as BMI and C-reactive protein.^24,38^ In AoU, AL comprised eight components routinely available in EHR data: blood pressure, BMI, glucose, hemoglobin, WBC, albumin, creatinine, and calcium. HbA1c, lipids, and C-reactive protein were considered but excluded because of high missingness. Laboratory measures were identified in the EHR using Logical Observation Identifiers Names and Codes (LOINC).

Each component was coded 1 if it fell outside the clinical reference range (above or below, as applicable) and 0 otherwise, using thresholds from clinical guidelines and laboratory references (Supplementary Table 4). The primary AL score was the sum of abnormal components (range 0-8), standardized to a z-score. Alternative scores were (i) a binary indicator of high AL (two or more abnormal components versus fewer than two); (ii) a factor-analytic score derived from exploratory factor analysis of the eight binary AL component indicators, which identified a single latent factor using standard minimum residual estimation; and (iii) the primary score restricted to participants with all eight components measured (complete case).

Missing AL components were imputed using random forests (50 trees per forest) as implemented in the missForest package (version 1.6.1)^60^ in R. The imputation model included all AL components and case status, age, sex, race and ethnicity, educational attainment, and household income.

### Outcomes

The primary outcome was exposome-related EoC, defined as the first diagnosis of malignant cancer before age 50 at head and neck (ICD-10 C00-C14), digestive (C15-C26), respiratory (C30-C39), soft tissue including mesothelioma (C45-C49), or urinary tract (C64-C68) sites. These sites were selected on the basis of established associations with environmental and lifestyle exposures, including tobacco, alcohol, diet, physical activity, and occupational exposures.^61–66^ The secondary outcome was all EoC, defined as the first diagnosis of malignant cancer before age 50, excluding breast cancer. Diagnoses were identified from OMOP condition concepts and grouped by ICD-10 site (Supplementary Table 6). This outcome included keratinocyte carcinomas (C44) and ill-defined, secondary, and unspecified malignant neoplasms (C76-C80). Fifty-three diagnoses were recorded with nonspecific concepts, such as primary malignant neoplasm or malignant neoplastic disease, that could not be assigned to a site; these were retained in the all-EoC outcome as unmapped, and a small number of them (20 or fewer) were recorded as carcinoma in situ. For exposome-related EoC, the 298 cases were compared with all 6,040 controls, which had been matched to the full case series.

AoU ascertains cancer outcomes primarily through linked EHR data mapped to the Observational Medical Outcomes Partnership common data model, supplemented by self-reported medical history. Baseline cancer-free status was confirmed using diagnosis codes before enrollment and survey responses on prior cancer history, and the first qualifying diagnosis after enrollment defined the event date.

### Statistical analysis

#### Exposome-AL association

Linear regression was used to estimate the association between the ERS and the standardized AL score, with sex-stratified models.

#### AL-EoC association

Logistic regression was used to estimate the association between AL and EoC, adjusted for age, sex, race and ethnicity, income, and education. Exploratory models were fitted separately for each cancer type; types with 20 or fewer cases were not reported, in accordance with the AoU Data and Statistics Dissemination Policy. Unlike the mediation models, these models were adjusted for covariates, so the odds ratios in Table 2 differ from the unadjusted outcome model used in the mediation analysis.

#### Mediation analysis

We used the mediation package in R (version 4.5.1),^67^ which implements a counterfactual-based framework comparing scenarios in which the mediator is held fixed or allowed to vary. The mediator model was a linear regression of AL on the ERS, and the outcome model was a logistic regression of EoC on AL and the ERS. Both models were fit in all participants in the analysis and included no additional covariates: cases and controls were matched on age, sex, and sociodemographic characteristics, and household income is a component of the ERS, so adjusting for it would remove part of the exposure. Adding age to both models yielded similar estimates (Supplementary Table 8). The contrast compared ERS values 1 SD above versus 1 SD below the sample mean. The indirect effect (average causal mediation effect) represents the portion of the ERS-EoC association operating through AL, and the direct effect (average direct effect) the remainder. Effects are expressed as differences in the predicted probability of case status within the analytic sample; because cases made up about 5% of the exposome-related analysis and 20% of the all-EoC analysis, effects on this scale are not directly comparable between outcomes. Uncertainty was estimated using 1,000 nonparametric bootstrap resamples. Interpretation of these effects as causal requires sequential ignorability, which cannot be verified empirically: no unmeasured confounding of the exposure-outcome, mediator-outcome, and exposure-mediator relationships, and no mediator-outcome confounders affected by the exposure.

#### Sensitivity analyses

We repeated the mediation analyses (i) using the binary and factor-analytic AL scores; (ii) restricted to participants with complete AL data; (iii) restricted to cases diagnosed at least 2 years after enrollment, to lengthen the interval between exposome assessment and diagnosis; (iv) adding age as a covariate to both models; and (v) stratified by sex. We assessed nonlinearity in the ERS-AL and AL-EoC associations using generalized additive models (mgcv), and repeated the mediation analysis with a generalized additive mediator model. Statistical significance was assessed at α=0.05.

#### Software

Data querying and matching were performed in Python (version 3.10.16). Statistical analyses were conducted in R (version 4.5.3).

## Data Availability

All data produced are available online via the NIH All of Us Workspace

## Ethics statement

The All of Us Research Program is conducted under a protocol approved by the All of Us Institutional Review Board, and all participants provided informed consent.^50^ This analysis used de-identified data accessed through the All of Us Researcher Workbench under the program’s data use and registration agreement. The All of Us Institutional Review Board has determined that research using these de-identified data does not constitute human subjects research; therefore, separate institutional review board approval was not required for this secondary analysis.

## Clinical trial registration

Not applicable. This was an observational case-control study of existing data; no intervention was assigned, and no clinical trial was conducted.

## Data availability

Data from the All of Us Research Program are available to registered researchers through the Researcher Workbench (https://www.researchallofus.org) after completion of required training and agreement to the data use policies. In accordance with these policies, the authors cannot share individual-level data.

## Code availability

Analysis code is available from the corresponding author upon reasonable request.

## Acknowledgments

We gratefully acknowledge All of Us participants for their contributions, without whom this research would not have been possible. We also thank the National Institutes of Health’s All of Us Research Program for making available the participant data examined in this study.

## Notes

### Competing Interest Statement

The authors have declared no competing interest.

### Author Declarations

The study used only openly available human data available through the NIH All of Us program.

