## Supplemental Tables and Figures for "Allostatic load as a potential pathway linking exposomic risk to early-onset cancer: a matched case-control study in the All of Us Research Program"

Supplementary Information

**Supplementary Figure 1. Temporal framework**

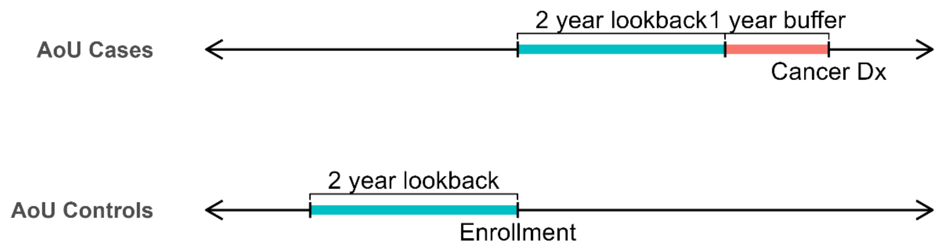


Exposome measures were obtained from surveys completed at enrollment. For controls, AL components were obtained from EHR data recorded within the 2 years before enrollment (consent to EHR sharing). For cases, AL components were obtained from EHR data recorded 1-3 years before cancer diagnosis, leaving at least a 1-year interval before diagnosis. Within each window, the most recent value of each component was used. Participants with cancer diagnosed before enrollment were excluded.

**Supplementary Figure 2. Nonlinearity of the exposome-allostatic load and allostatic load-early-onset cancer associations**

**a**

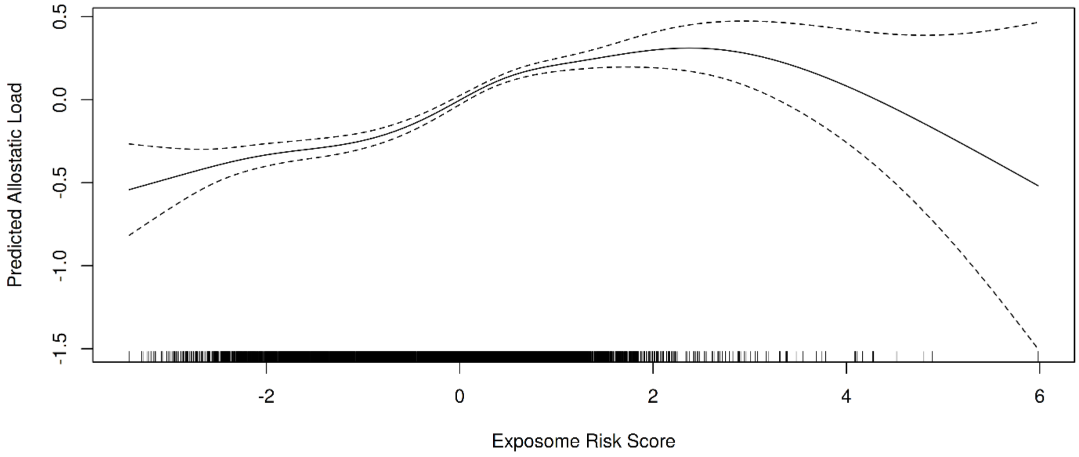


**b**

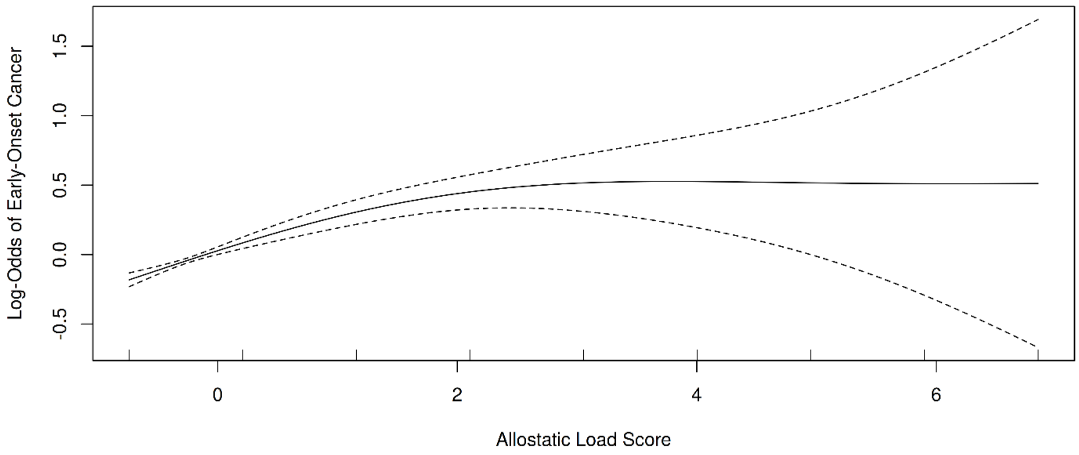


Generalized additive model fits (solid lines) with 95% confidence intervals (dashed lines). (a) Standardized AL as a function of the exposomic risk score. (b) Log-odds of early-onset cancer as a function of the standardized AL score. Rug plots show observed values.

**Supplementary Table 1. Selected mechanisms linking allostatic load to carcinogenesis**

| Category | Mechanism |
|---|---|
| Chronic inflammation[46] | Sustained inflammatory signaling activates NF-κB pathways promoting cell proliferation and angiogenesis; elevated pro-inflammatory cytokines (IL-6, TNF-α, CRP) create tumor-promoting microenvironments |
| Metabolic dysfunction[68] | Dysregulated glucose metabolism and insulin resistance alter cellular energetics favoring carcinogenesis; adipose tissue dysfunction releases adipokines that promote tumor growth and metastasis |
| Immune dysfunction[45] | Impaired natural killer cell cytotoxicity reduces tumor surveillance; chronic stress hormones suppress T-cell proliferation and shift immune responses toward immunosuppression |
| Genomic instability[69] | Oxidative stress induces DNA damage while impairing repair mechanisms; accelerated telomere shortening and epigenetic aging increase somatic mutation burden |

**Supplementary Table 2. Selected studies of allostatic load and cancer outcomes**

| Study | Population | Operationalization of AL | Association |
|---|---|---|---|
| Zhao et al. (2025)[34] | 304,959 UK Biobank adults (incidence); 1,794 patients with colorectal cancer (mortality) | 12 biomarkers across 4 systems: metabolic (HbA1c, HDL, LDL, total cholesterol, triglycerides, waist-to-hip ratio); cardiovascular (systolic and diastolic blood pressure, pulse rate); inflammatory/immune (CRP, IGF-1); renal (creatinine) | Colorectal cancer incidence: 43% higher risk, highest vs lowest AL quartile; 39.2% of cases attributable to AL (95% CI 36.2-42.3%). All-cause mortality: HR 1.71 (95% CI 1.16-2.50), highest vs lowest quartile |
| Akinyemiju et al. (2020)[32] | 29,701 adults aged ≥45 years | Sum score of 10 biomarkers (albumin, CRP, HDL, total cholesterol, heart rate, systolic and diastolic blood pressure, serum creatinine, blood urea nitrogen, waist circumference) | Cancer-specific mortality: HR 1.07 (95% CI 1.03-1.12) |
| Obeng-Gyasi et al. (2023)[33] | 4,459 patients with stage I-III breast cancer | 10 biomarkers across 4 systems: cardiovascular (heart rate, systolic and diastolic blood pressure); metabolic (BMI, alkaline phosphatase, glucose, albumin); renal (creatinine, blood urea nitrogen); immune (WBC); quartile method | All-cause mortality: HR 1.46 (95% CI 1.11-1.93) |

BMI, body mass index; CRP, C-reactive protein; HDL, high-density lipoprotein; HR, hazard ratio; IGF-1, insulin-like growth factor 1; LDL, low-density lipoprotein; WBC, white blood cell count.

**Supplementary Table 3. Exposome variables used to derive the exposomic risk score**

| Variable | Description |
|---|---|
| PANES Walk & Bike score | Validated scale assessing neighborhood walkability and access to recreation facilities[70] |
| PANES Crime & Safety score | Validated scale assessing neighborhood crime and safety[70] |
| Everyday Discrimination score | Validated scale assessing perceived discrimination in everyday life[71] |
| Discrimination in Health Care Settings score | Validated scale assessing perceived discrimination in health care settings[72] |
| Social Support score | Validated scale assessing social support[73] |
| Social Cohesion score | Validated scale assessing perceived neighborhood cohesion[74] |
| Neighborhood Social Disorder score | Validated scale assessing neighborhood social order and disorder[75] |
| Neighborhood Physical Disorder score | Validated scale assessing neighborhood physical order and disorder[75] |
| Household size | “Not including yourself, how many other people live at home with you?” |
| Household income | “What is your annual household income from all sources?” |
| Number of other substances used | Count of substances selected for “In your LIFETIME, which of the following substances have you ever used?” |
| Housing instability | “In the last 12 months, how many times have you or your family moved from one home to another?” |

Multi-item scales were derived from validated instruments implemented in AoU and documented in the Social Determinants of Health data dictionary; the remaining variables were single-item survey questions. Nine binary items were excluded from the factor analysis but retained as predictors in the imputation model: born in the USA, public insurance, home ownership, ≥100 lifetime cigarettes, smokeless tobacco use, weekly alcohol use, rural-related delayed care, travel outside the US, food insecurity. PANES, Physical Activity Neighborhood Environment Survey.

**Supplementary Table 4. Thresholds for abnormal allostatic load components**

| Component | Abnormal range | Units | Source |
|---|---|---|---|
| Blood pressure | Systolic >140 or diastolic >90 | mmHg | Williams et al. 2018 (ESC/ESH)[76] |
| Body mass index | <18.5 or >35 | kg/m² | Jensen et al. 2014 (AHA/ACC/TOS)[77] |
| Glucose | >140 | mg/dL | Moghissi et al. 2009[78] |
| Hemoglobin | Men: <109 or >180; women: <109 or >160 | g/L | El Brihi & Pathak 2024[79] |
| White blood cell count | <4 or >11 | ×10⁹/L | El Brihi & Pathak 2024[79] |
| Albumin | <35 | g/L | Gatta et al. 2012[80] |
| Creatinine | Men: >1.3; women: >1.1 | mg/dL | Abcar et al. 2004[81] |
| Calcium | <8.6 or >10.0 | mg/dL | Mayo Clinic Laboratories[82] |

Each component was coded 1 if its value fell strictly beyond the listed threshold and 0 otherwise.

**Supplementary Table 5. Missingness and abnormal values by allostatic load component**

| Component | Missing (%) | Abnormal, controls (%) | Abnormal, cases (%) | Abnormal, overall (%) |
|---|---|---|---|---|
| Creatinine | 48.00 | 5.04 | 6.85 | 5.50 |
| Hemoglobin | 47.03 | 12.53 | 15.19 | 13.20 |
| Blood pressure | 24.86 | 9.92 | 15.23 | 10.98 |
| Body mass index | 10.19 | 27.95 | 30.38 | 28.40 |
| Glucose | 47.67 | 9.36 | 9.88 | 9.49 |
| White blood cell count | 50.26 | 18.71 | 18.76 | 18.72 |
| Albumin | 56.86 | 12.48 | 12.30 | 12.43 |
| Calcium | 49.97 | 13.50 | 14.34 | 13.71 |

Percentages of abnormal values are among participants with an observed value for each component.

**Supplementary Table 6. Cancer group definitions**

| Cancer group | ICD-10 codes | Exposome-related (primary outcome) |
|---|---|---|
| Head and neck | C00-C14 | Yes |
| Digestive | C15-C26 | Yes |
| Respiratory | C30-C39 | Yes |
| Soft tissue and mesothelioma | C45-C49 | Yes |
| Urinary tract | C64-C68 | Yes |
| Bone and articular cartilage | C40-C41 | No |
| Skin | C43-C44 | No |
| Female genital | C51-C58 | No |
| Male genital | C60-C63 | No |
| Eye, brain, and central nervous system | C69-C72 | No |
| Endocrine | C73-C75 | No |
| Neuroendocrine | C7A | No |
| Other and ill-defined | C76-C80 | No |
| Hematologic and lymphoid | C81-C96 | No |
| Unmapped | Nonspecific malignant neoplasm concepts without an assignable site | No |

Diagnoses were identified from OMOP condition concepts and grouped by ICD-10 site. Breast cancer (C50) was excluded from all analyses (Methods). The skin group includes keratinocyte carcinomas (C44). A small number of unmapped diagnoses (20 or fewer) were recorded as carcinoma in situ. ICD-10, International Classification of Diseases, Tenth Revision.

**Supplementary Table 7. Exploratory associations between allostatic load and early-onset cancer by cancer type**

| Cancer type | Cases | Odds ratio (95% CI) | p |
|---|---|---|---|
| **Exposome-related cancers** | | | |
| Digestive | 134 | 1.24 (1.07-1.44) | 0.004 |
| Urinary tract | 72 | 1.50 (1.26-1.77) | <0.001 |
| Respiratory | 41 | 1.18 (0.90-1.55) | 0.233 |
| Head and neck | 27 | 1.45 (1.10-1.91) | 0.009 |
| Soft tissue and mesothelioma | 24 | 1.21 (0.85-1.71) | 0.288 |
| **Other cancers** | | | |
| Hematologic and lymphoid | 377 | 1.46 (1.34-1.58) | <0.001 |
| Female genital | 270 | 1.16 (1.04-1.30) | 0.007 |
| Skin | 199 | 0.83 (0.70-0.99) | 0.034 |
| Endocrine | 145 | 1.15 (0.99-1.34) | 0.061 |
| Bone and articular cartilage | 73 | 0.78 (0.59-1.05) | 0.098 |
| Eye, brain, and central nervous system | 51 | 0.89 (0.65-1.21) | 0.458 |
| Male genital | 29 | 1.21 (0.89-1.66) | 0.228 |
| Unmapped | 53 | 1.18 (0.93-1.50) | 0.181 |
| Other and ill-defined | ≤20 | Not reported | |
| Neuroendocrine | ≤20 | Not reported | |

Odds ratios per 1-SD increase in the primary AL score from logistic regression adjusted for age, sex, race and ethnicity, income, and education. Cancer types with 20 or fewer cases are not reported, per the AoU Data and Statistics Dissemination Policy. These analyses are exploratory and were not adjusted for multiple comparisons.

**Supplementary Table 8. Sensitivity analyses for mediation of the exposome-early-onset cancer association by allostatic load**

| Analysis and outcome | Total effect | Direct effect | Indirect effect | Proportion mediated |
|---|---|---|---|---|
| **Cases diagnosed at least 2 years after enrollment** | | | | |
| Exposome-related EoC | 0.008 (0.002, 0.015); p=0.010 | 0.006 (0.001, 0.013); p=0.026 | 0.001 (0.001, 0.002); p<0.001 | 16.4% (5.2%, 64.8%); p=0.010 |
| All EoC | 0.020 (0.008, 0.032); p=0.004 | 0.015 (0.003, 0.027); p=0.016 | 0.004 (0.002, 0.006); p<0.001 | 21.8% (9.0%, 61.0%); p=0.004 |
| **Additional age adjustment** | | | | |
| Exposome-related EoC | 0.019 (0.010, 0.029); p<0.001 | 0.015 (0.005, 0.024); p=0.004 | 0.003 (0.002, 0.004); p<0.001 | 16.0% (7.5%, 37.9%); p<0.001 |
| All EoC | 0.022 (0.003, 0.040); p=0.016 | 0.010 (−0.007, 0.028); p=0.236 | 0.011 (0.008, 0.014); p<0.001 | 50.5% (24.8%, 229.9%); p=0.016 |
| **Factor-analytic AL score** | | | | |
| Exposome-related EoC | 0.018 (0.007, 0.028); p<0.001 | 0.015 (0.004, 0.025); p=0.004 | 0.002 (0.001, 0.003); p<0.001 | 12.0% (5.7%, 36.9%); p<0.001 |
| All EoC | 0.022 (0.004, 0.040); p=0.014 | 0.014 (−0.003, 0.033); p=0.120 | 0.007 (0.005, 0.010); p<0.001 | 33.9% (14.6%, 142.5%); p=0.014 |
| **Binary AL score (≥2 abnormal components)** | | | | |
| Exposome-related EoC | 0.019 (0.010, 0.029); p<0.001 | 0.015 (0.005, 0.025); p=0.002 | 0.003 (0.001, 0.005); p<0.001 | 14.8% (6.0%, 39.3%); p<0.001 |
| All EoC | 0.023 (0.005, 0.043); p=0.010 | 0.014 (−0.004, 0.034); p=0.148 | 0.009 (0.005, 0.013); p<0.001 | 39.0% (16.5%, 148.7%); p=0.010 |
| **Male participants** | | | | |
| Exposome-related EoC | 0.041 (0.014, 0.068); p=0.004 | 0.034 (0.008, 0.060); p=0.014 | 0.005 (0.002, 0.008); p<0.001 | 11.5% (4.2%, 42.2%); p=0.004 |
| All EoC | 0.005 (−0.027, | −0.005 (−0.037, | 0.011 (0.005, 0.016); p<0.001 | Not estimable† |

| Analysis and outcome | Total effect | Direct effect | Indirect effect | Proportion mediated |
|---|---|---|---|---|
| | 0.040); p=0.782 | 0.029); p=0.744 | | |
| **Female participants** | | | | |
| Exposome-related EoC | 0.008 (−0.001, 0.017); p=0.114 | 0.005 (−0.004, 0.014); p=0.310 | 0.003 (0.001, 0.004); p<0.001 | Not estimable† |
| All EoC | 0.027 (0.008, 0.048); p=0.012 | 0.016 (−0.003, 0.038); p=0.094 | 0.010 (0.006, 0.014); p<0.001 | 37.7% (16.8%, 120.1%); p=0.012 |
| **Complete AL data** | | | | |
| Exposome-related EoC | 0.028 (0.005, 0.054); p=0.018 | 0.025 (0.001, 0.052); p=0.046 | 0.002 (−0.002, 0.006); p=0.380 | 6.4% (−13.8%, 47.6%); p=0.398 |
| All EoC | 0.042 (0.004, 0.081); p=0.022 | 0.037 (0.000, 0.077); p=0.052 | 0.004 (−0.003, 0.012); p=0.282 | 10.3% (−15.5%, 75.5%); p=0.292 |
| **Generalized additive mediator model** | | | | |
| Exposome-related EoC | 0.019 (0.008, 0.029); p=0.002 | 0.014 (0.004, 0.024); p=0.006 | 0.005 (0.003, 0.006); p<0.001 | 24.1% (13.3%, 54.0%); p=0.002 |
| All EoC | 0.024 (0.006, 0.041); p=0.014 | 0.011 (−0.008, 0.028); p=0.244 | 0.014 (0.009, 0.018); p<0.001 | 55.7% (28.7%, 209.4%); p=0.014 |

All models were fit without covariates except the age-adjusted analysis, which added age to both models. The generalized additive analysis replaced the linear mediator model with a generalized additive model. Effects are differences in the predicted probability of case status comparing an ERS 1 SD above versus 1 SD below the mean; values in parentheses are 95% confidence intervals from 1,000 nonparametric bootstrap resamples. †Not estimable because the total effect was not statistically significant; for all EoC among male participants, the estimated proportion mediated was 205.1% (95% CI −948.7% to 808.0%).
